# Impact of vaccination on COVID-19-associated admissions to critical care in England: a population cohort study of linked data

**DOI:** 10.1101/2022.10.03.22280649

**Authors:** David A Harrison, Peter J Watkinson, James C Doidge, Manu Shankar-Hari, Paul R Mouncey, Martina Patone, Carol A C Coupland, Julia Hippisley-Cox, Kathryn M Rowan

## Abstract

**Introduction:** This study aims to explore the impact of COVID-19 vaccination on critical care by examining associations between vaccination and admission to critical care with COVID-19 during England’s Delta wave, by age group, dose, and over time.

**Methods:** We used linked routinely-collected data to conduct a population cohort study of patients admitted to adult critical care in England for management of COVID-19 between 1 May and 15 December 2021. Included participants were the whole population of England aged 18 years or over (44.7 million), including 10,141 patients admitted to critical care with COVID-19. The intervention was vaccination with one, two, or a booster/three doses of any COVID-19 vaccine.

**Results:** Compared with unvaccinated patients, vaccinated patients were older (median 64 years for patients receiving two or more doses versus 50 years for unvaccinated), with higher levels of severe comorbidity (20.3% versus 3.9%) and immunocompromise (15.0% versus 2.3%). Compared with patients who were unvaccinated, those vaccinated with two doses had a relative risk reduction (RRR) of between 90.1% (patients aged 18–29, 95% CI, 86.8% to 92.7%) and 95.9% (patients aged 60–69, 95% CI, 95.5% to 96.2%). Waning was only observed for those aged 70+, for whom the RRR reduced from 97.3% (91.0% to 99.2%) to 86.7% (85.3% to 90.1%) between May and December but increased again to 98.3% (97.6% to 98.8%) with a booster/third dose.

**Conclusion:** Important demographic and clinical differences exist between vaccinated and unvaccinated patients admitted to critical care with COVID-19. While not a causal analysis, our findings are consistent with a substantial and sustained impact of vaccination on reducing admissions to critical care during England’s Delta wave, with evidence of waning predominantly restricted to those aged 70+.

## Introduction

The COVID-19 pandemic has seen critical care services overwhelmed in many parts of the world.(1-3) The demand that COVID-19 has placed on critical care services has been a core driver of policy responses to the pandemic. On 8 December 2020, the United Kingdom (UK) commenced its COVID-19 vaccination programme,(4) at which time over 7000 patients with COVID-19 had already been admitted for critical care across England, Wales and Northern Ireland.(5) By 15 December 2021, 4.42 billion people globally had received a vaccine against COVID-19, including over 51 million in the UK.(6) The UK vaccination programme prioritised older care home residents and older adults and then rolled out in order of age and clinical risk factors. In line with the UK vaccination prioritisation, age has been identified as the strongest predictor of COVID-19 mortality with older patients significantly more likely to die during their admission for critical care.(7) In the context of waning of vaccine-acquired antibodies and immune escape by variants, understanding the extent to which COVID-19 vaccines protect against critical illness and their impact on admission to critical care is crucial for service planning and policy development.

Randomised controlled trials of vaccine efficacy inevitably have relatively low numbers of critical care admissions,(8-10) but recent population studies suggest that vaccination reduces the risk of admission for critical care.(11-13) However, despite widespread vaccination, substantial numbers of people are still being admitted to critical care units for the management of COVID-19 internationally, with unvaccinated patients reported to be overrepresented.(14, 15)

Using routinely collected data, our aim was to explore how vaccinated and unvaccinated patients contributed to critical care use. We compare patient characteristics by vaccination status, the proportion of patients admitted for critical care with each vaccination status to proportions in the general population by month, and estimate rates of admission by age and vaccination status over time. Finally, we explore any association between vaccination status and admission rate.

## Methods

### Study design

Population cohort study using routinely collected healthcare data.

### Ethical approval

NHS Research Ethics Committee approval was obtained from East Midlands-Derby Research Ethics Committee (ref 04/03/2021).

### Data sources

The Intensive Care National Audit & Research Centre (ICNARC) Case Mix Programme is the national clinical audit for adult critical care in England, Wales and Northern Ireland. All adult general critical care units providing Level 3 critical care (intensive care) participate in the audit and submit data on consecutive admissions. Some specialist critical care units and Level 2 (high dependency only) units also participate. During the COVID-19 pandemic, coverage was widened to include many temporary critical care areas (‘surge areas’) providing critical care to patients with COVID-19.

Data for SARS-CoV-2 positive test results were extracted from the Second Generation Surveillance System (SGSS), maintained by the UK Health Security Agency.(16) Data for COVID-19 vaccinations were extracted from the National Immunisation Management System (NIMS), maintained by the South, Central and West Commissioning Unit on behalf of NHS England and NHS Improvement.(17) The NIMS database includes vaccine type, date and doses for all people vaccinated in England. The SGSS and NIMS data are collated, maintained and quality assured by NHS Digital.

Numbers of patients vaccinated by age and date were taken from the UK Government Coronavirus Dashboard.(18) Population denominators were taken from Office for National Statistics (ONS) population estimates from the 2021 census.(19)

### Data linkage

Data linkage was undertaken on the QResearch Trusted Research Environment (TRE) hosted at the University of Oxford. Data were linked using a pseudonymised version of the patient’s NHS number (a unique number assigned to all patients receiving care in the UK National Health Service). The NHS number was encrypted prior to transfer to QResearch with a one-way hashing algorithm using OpenPseudonymiser v2.0.2c (University of Nottingham).

### Inclusion criteria

Patients were included in the analysis if they were: (1) admitted to a critical care unit in England participating in the ICNARC Case Mix Programme with confirmed COVID-19 recorded as the primary, secondary or ultimate primary reason for admission to the critical care unit between 1 May 2021 (corresponding to the start of the Delta variant wave of the pandemic in England) and 15 December 2021 (the latest date for which vaccination data were available at the point of analysis); (2) aged 18 years or over on the date of admission to critical care; and (3) successfully linked to a positive SARS-CoV-2 test result in a time window from 28 days prior to admission to the critical care unit to 2 days after admission to the critical care unit.

The following patients were excluded from the analysis: patients with a missing or invalid NHS number (precluding linkage with other data sources); and patients not usually resident in England. Where there were multiple critical care admissions of the same patient in the study period, only the first admission was included in the analysis.

### Exposure definitions

COVID-19 vaccination status was based on administration of any UK approved COVID-19 vaccine – specifically, ChAdOx1 (Oxford/Astra-Zeneca), BNT162b2 (Pfizer/BioNTech) or mRNA-1273 (Moderna). Based on published knowledge of the delay between vaccination and effectiveness,(20) vaccination status was assessed at 14 days prior to the positive SARS-CoV-2 test and categorised as:

- Unvaccinated: Either no linked vaccination record in NIMS or first dose of vaccine received within 14 days prior to the positive SARS-CoV-2 test
- One dose: First dose of vaccine received at least 14 days prior to the positive SARS-CoV-2 test (includes patients that had received a second dose within 14 days prior to the positive SARS-CoV-2 test)
- Two doses: Second dose of vaccine received at least 14 days prior to the positive SARS-CoV-2 test (includes patients that had received a booster or third dose within 14 days prior to the positive SARS-CoV-2 test)
- Booster or three doses: Booster or third dose of vaccine received at least 14 days prior to the positive SARS-CoV-2 test

To test the sensitivity of results to the choice of the 14-day lag for assessing vaccination status, analyses were repeated based on the vaccination status on the date of the positive SARS-CoV-2 test.

### Statistical analysis

The following characteristics of patients critically ill with confirmed COVID-19 were summarised by vaccination status over the whole time period:

- Age at admission to critical care
- Sex (male or female)
- Ethnicity – recorded using NHS Ethnic Category codes (based on UK 2011 census) and combined for reporting as White, Mixed, Asian, Black or Other
- Quintile of Index of Multiple Deprivation
- Any dependency prior to the onset of acute illness resulting in admission to hospital – assessed as the requirement for assistance with activities of daily living (washing, dressing, etc.)
- Any severe comorbidity – evident within the six months prior to admission to critical care and documented at or prior to admission to critical care, as recorded for the Acute Physiology And Chronic Health Evaluation (APACHE) II:

○ Cardiovascular (symptoms at rest; New York Heart Association functional class IV)
○ Respiratory (shortness of breath with light activity or receipt of home ventilation)
○ Renal (renal replacement therapy for end-stage renal disease)
○ Liver (biopsy-proven cirrhosis, portal hypertension or hepatic encephalopathy)
○ Immunocompromised (radiotherapy, chemotherapy, daily high-dose steroid treatment, metastatic disease, haematological malignancy, HIV/AIDS, congenital immunohumoral or cellular immune deficiency state)
- Immunocompromised – defined as above
- Body mass index – calculated from weight and height, measured or estimated on admission to critical care
- Pregnancy status for women aged 18-49 – currently pregnant on admission to critical care, recently pregnant (within the previous 6 weeks) or not known to be pregnant
- COVID-19 reported as the primary (including ultimate primary), rather than secondary, reason for admission to critical care
- Invasive ventilation at any time during the first 24 hours following admission to critical care
- The ratio of partial pressure of oxygen in arterial blood to fraction of inspired oxygen (PaO_2_/FiO_2_ ratio) and corresponding FiO_2_ from the arterial blood gas with the lowest PaO_2_ during the first 24 hours following admission to critical care

The proportion of patients within each vaccination status category was summarised by month of admission to critical care and compared with the proportion of patients with each vaccination status in the population of England (averaged over the period 14 days prior to the month of admission to critical care).

Rates of critical care admission overall, by age group (18-29 years, 30-39 years, 40-49 years, 50-59 years, 60-69 years and 70 years and over), and by age group and vaccination status were calculated as the number of critical care admissions divided by the total exposure in person-days. The exposure for each patient was calculated as the size of the population of England with the corresponding vaccination status at 14 days prior to the date of admission to critical care. The size of the vaccinated populations in England was based on NIMS data, via the UK Government COVID-19 dashboard. The size of the unvaccinated population was estimated by subtracting the numbers that had received one or more doses of vaccine from the ONS 2021 population estimates. Rates were calculated using a 30-day moving average. In line with the data sharing agreement governing access to the data, results are not reported for any 30-day period during which five or fewer patients were admitted to critical care.

The association between vaccination and reduced admission rates was reported as the relative risk reduction (RRR), calculated as one minus the incidence rate ratio,(21) for each vaccinated group compared with the unvaccinated group. This was plotted as a 30-day moving average by age group.

## Results

Of 11,431 patients admitted to adult critical care units in England with COVID-19 between 1 May 2021 and 15 December 2021, 190 (1.7%) were missing NHS number and unable to be linked, 68 (0.6%) were aged under 18 years and 60 (0.5%) were not resident in England. Of the remaining 11,113 patients, 10,141 (91.3%) were successfully linked to a positive SARS-CoV-2 test result between 28 days prior to admission to the critical care unit and 2 days after admission to the critical care unit.

Linkage with NIMS records showed 419 (4.1%) patients had received one dose of vaccine at least 14 days prior to the positive SARS-CoV-2 test, 3615 (35.6%) had received two doses and 152 (1.5%) had received a booster or three doses. The remaining 5955 (58.7%) were classified as unvaccinated, including 113 (1.1%) that had received a first dose of vaccine within the 14 days prior to the positive SARS-CoV-2 test. Of 11,224 doses of vaccine received, 6048 (53.9%) were ChAdOx1 (Oxford/Astra-Zeneca), 4849 (43.2%) were BNT162b2 (Pfizer/BioNTech) and 327 (2.9%) were mRNA-1273 (Moderna).

### Patient characteristics by vaccination status

Characteristics of patients admitted to critical care by vaccination status are reported in Table 1. Compared with critical care patients who had received two or more doses of vaccine, unvaccinated patients were younger, more likely to be female, more likely to be of a non-White ethnicity, more likely to be resident in a deprived area, and less likely to have any dependency prior to their acute illness or to have any severe comorbidity (particularly immunocompromise). Over one-third of unvaccinated women aged 18-49 years at admission to critical care (467/1282) were either currently pregnant or recently pregnant (within 6 weeks prior to admission to critical care). In contrast, only five or fewer vaccinated women were admitted to critical care that were either currently or recently pregnant (the threshold required for reporting precise numbers). While the vast majority of patients across all categories had COVID-19 reported as their primary reason for admission to critical care, this was slightly higher for unvaccinated than for vaccinated patients (96% versus 92%). A similar proportion of unvaccinated and vaccinated patients were invasively ventilated during the first 24 hours following admission to the critical care unit (21% versus 20%), and there was little difference in either median PaO_2_/FiO_2_ ratio or FiO_2_ received.

**Table 1.** Characteristics of patients admitted to critical care with COVID-19 between May and December 2021 in England by vaccination status.

|  | Vaccination status <sup>a</sup> |  |  |
| --- | --- | --- | --- |
|  | Unvaccinated<br>[N=5955] | One dose<br>[N=419] | Two or more doses <sup>b</sup><br>[N=3767] |
| Age (years), median (IQR) | 50 (38, 60) | 52 (44, 63) | 64 (55, 72) |
| Age group, n (%) |  |  |  |
| 18-29 | 588 (9.9) | 17 (4.1) | 52 (1.4) |
| 30-39 | 1094 (18.4) | 49 (11.7) | 151 (4.0) |
| 40-49 | 1270 (21.3) | 102 (24.3) | 399 (10.6) |
| 50-59 | 1400 (23.5) | 116 (27.7) | 815 (21.6) |
| 60-69 | 1075 (18.1) | 98 (23.4) | 1123 (29.8) |
| 70+ | 528 (8.9) | 37 (8.8) | 1227 (32.6) |
| Sex, n (%) |  |  |  |
| Female | 2444 (41.0) | 165 (39.4) | 1265 (33.6) |
| Male | 3511 (59.0) | 254 (60.6) | 2502 (66.4) |
| Ethnicity, n (%) | [N=5477] | [N=395] | [N=3565] |
| White | 3580 (65.4) | 267 (67.6) | 3045 (85.4) |
| Mixed | 121 (2.2) | 11 (2.8) | 27 (0.8) |
| Asian | 700 (12.8) | 75 (19.0) | 285 (8.0) |
| Black | 645 (11.8) | 26 (6.6) | 114 (3.2) |
| Other | 431 (7.9) | 16 (4.1) | 94 (2.6) |
| Quintile of IMD, n (%) | [N=5888] | [N=416] | [N=3732] |
| 1 (least deprived) | 460 (7.8) | 51 (12.3) | 570 (15.3) |
| 2 | 674 (11.4) | 50 (12.0) | 686 (18.4) |
| 3 | 1011 (17.2) | 57 (13.7) | 700 (18.8) |
| 4 | 1537 (26.1) | 99 (23.8) | 830 (22.2) |
| 5 (most deprived) | 2206 (37.5) | 159 (38.2) | 946 (25.3) |
| Any dependency, n/N (%) | 359/5522 (6.5) | 47/394 (11.9) | 564/3444 (16.4) |
| Any severe comorbidity, n/N (%) | 228/5781 (3.9) | 51/412 (12.4) | 744/3659 (20.3) |
| Immunocompromise, n/N (%) | 132/5781 (2.3) | 28/412 (6.8) | 549/3659 (15.0) |
| BMI (kg/m<sup>2</sup>), n (%) | [N=5542] | [N=392] | [N=3450] |
| <25 | 954 (17.2) | 73 (18.6) | 726 (21.0) |
| 25-29.9 | 1738 (31.4) | 106 (27.0) | 1034 (30.0) |
| 30-34.9 | 1315 (23.7) | 106 (27.0) | 808 (23.4) |
| 35-39.9 | 786 (14.2) | 46 (11.7) | 461 (13.4) |
| 40+ | 749 (13.5) | 61 (15.6) | 421 (12.2) |
| Pregnancy, n (% of females aged 18-49) | [N=1282] | [N=72] | [N=298] |
| Currently pregnant | 262 (20.4) | N≤5 | N≤5 |
| Recently pregnant (within 6 weeks) | 205 (16.0) | N≤5 | N≤5 |
| Not known to be pregnant | 815 (63.6) |  |  |
| COVID-19 reported as primary, rather than secondary, reason for admission to critical care, n (%) | 5720 (96.1) | 382 (91.2) | 3458 (91.8) |
| Invasively ventilated during the first 24h following admission to critical care, n/N (%) | 1187/5742 (20.7) | 79/413 (19.1) | 711/3599 (19.8) |
| Arterial blood gas measurements from the first 24h following admission to critical care <sup>c</sup>, median (IQR) | [N=5362] | [N=390] | [N=3324] |
| PaO<sub>2</sub>/FiO<sub>2</sub> ratio (kPa) | 13.2 (9.9, 18.0) | 13.7 (10.3, 19.0) | 13.6 (9.8, 19.3) |
| FiO<sub>2</sub> | 0.60 (0.45, 0.80) | 0.60 (0.40, 0.75) | 0.60 (0.40, 0.80) |

BMI, body mass index; IMD, Index of Multiple Deprivation; IQR, interquartile range; N, number of patients; N≤5 indicates results have been suppressed due to five or fewer patients in a category
<sup>a</sup> Vaccination status assessed at 14 days prior to the positive SARS-CoV-2 test
<sup>b</sup> Includes patients that received booster/third dose (N=152) due to small sample size
<sup>c</sup> From the arterial blood gas with the lowest PaO<sub>2</sub> during the first 24 hours following admission to critical care

### Vaccination status over time

Between May and December 2021, the proportion of the general population of England aged 18 years or older that were unvaccinated decreased from 36% to 10%. Among patients admitted to critical care, the proportion that were unvaccinated reduced from 75% in May 2021 to 47% in October 2021, before increasing again to 61% in December 2021 (Figure 1). With the rollout of the booster programme from September 2021 onwards, the proportion of the general population that had received a booster/third dose increased to 31% in December 2021. By contrast, only 9% of patients admitted to critical care in December 2021 had received a booster/third dose.

**Figure 1.**
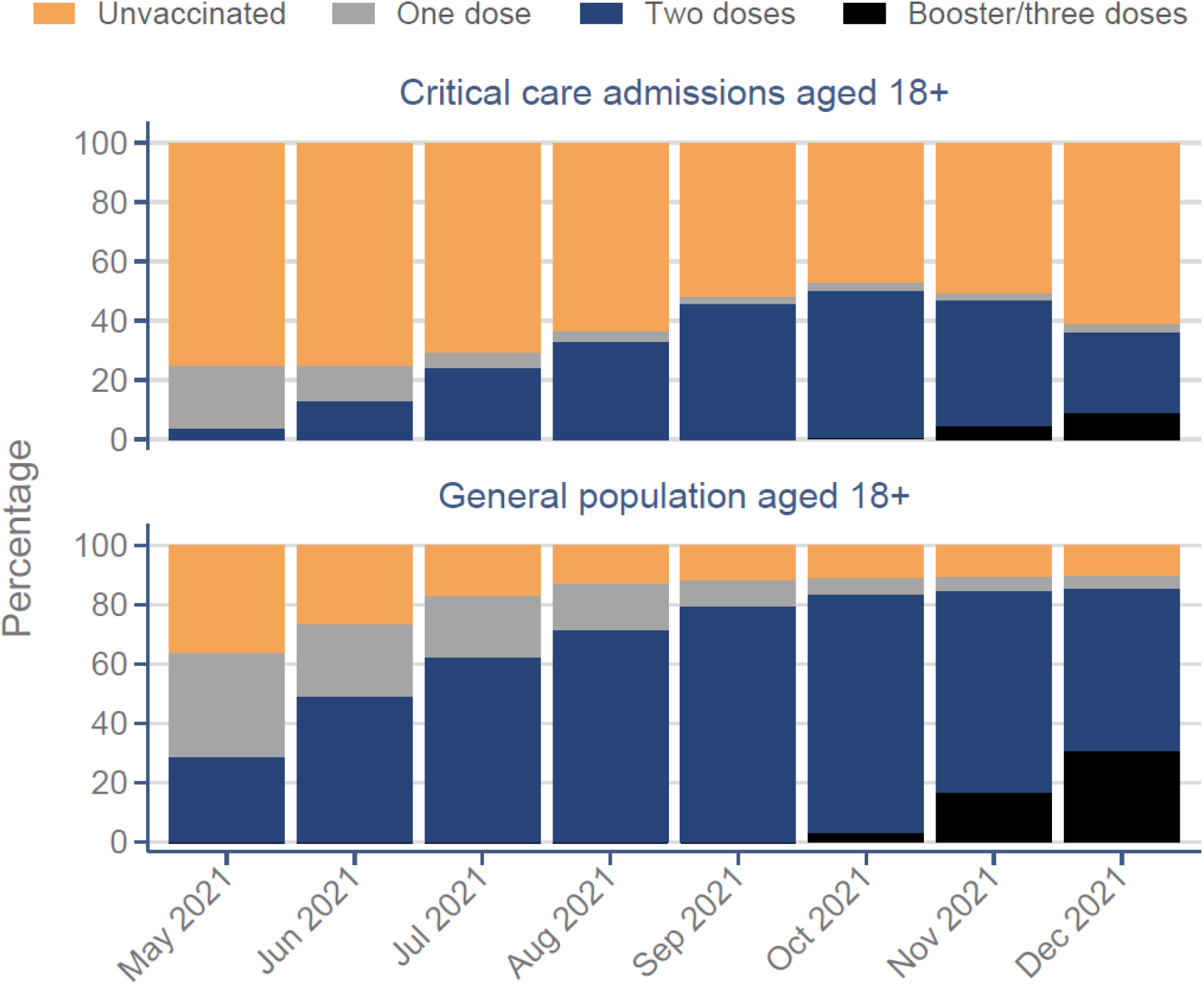
Vaccination status of critical care admissions compared with the general population by month.

### Rates of admission to critical care over time

From an initial low of 0.01 admissions per 100,000 population per day in May 2021, the rate of admissions to critical care in the overall population peaked at around 0.15 per 100,000 per day in July and August 2021 and at a similar level again in November 2021 (Supplementary appendix, Figure S1). Admission rates in the overall population varied four-fold by age group and peaked earlier, in mid to late July, for people aged 18-59 years compared with September for those aged 60 years and over (Supplementary appendix, Figure S2).

The rate of admission to critical care by age group and vaccination status is shown in Figure 2. Among the unvaccinated, admission rates varied substantially by age, peaking at 4.5 admissions per 100,000 population per day for 60–69-year-olds compared with 0.2 for 18–29-year-olds. Across all age groups, the admission rates for the unvaccinated were substantially greater than for all three categories of vaccinated patients.

**Figure 2.**
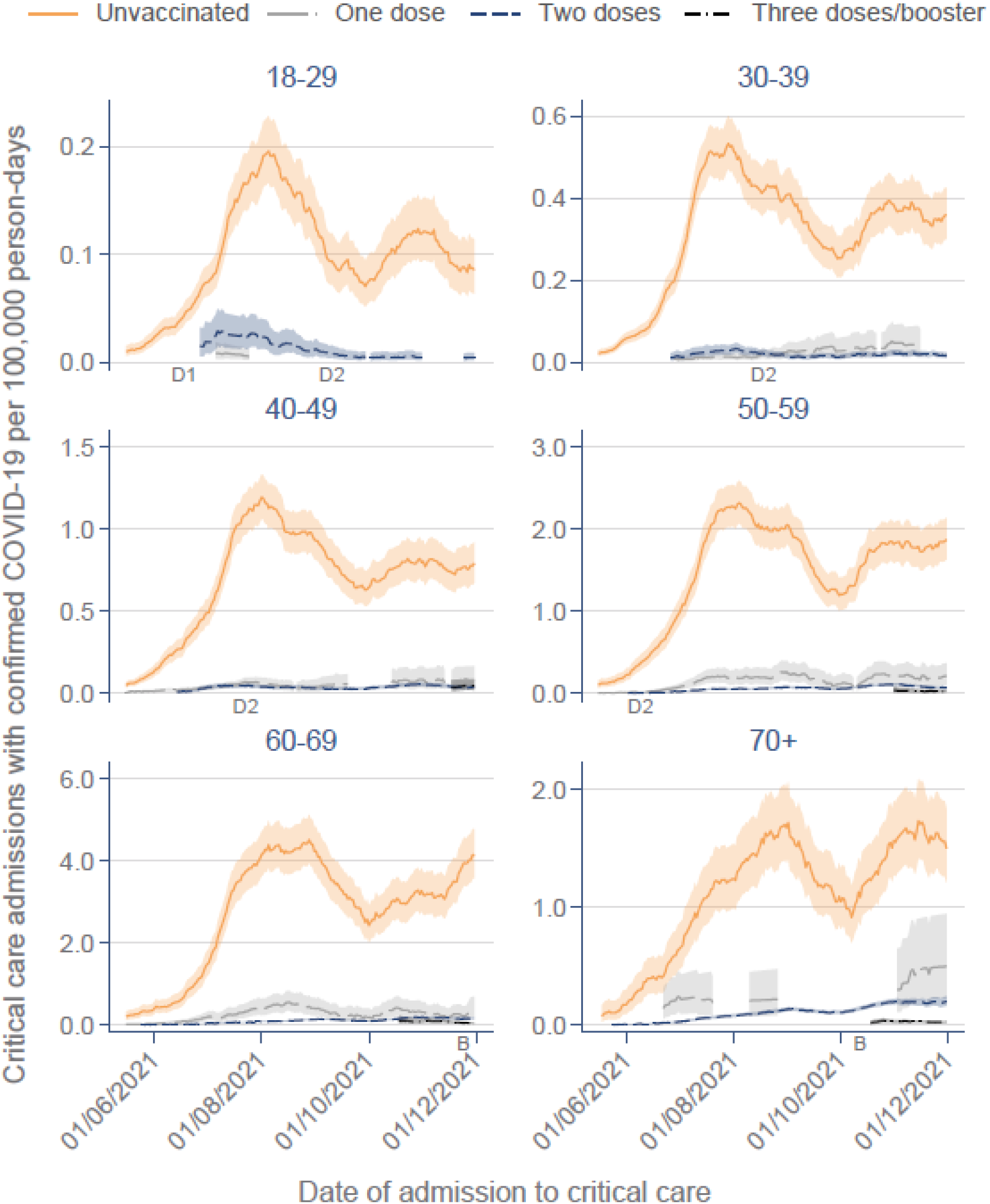
Rate of admission to critical care by age group and vaccination status over time. Vaccination status assessed at 14 days prior to the positive SARS-CoV-2 test. 30-day moving average. D1=date on which dose 1 of vaccine was offered to all people within the age group (predates the analysis time period for age groups of 30 years and over). D2=date on which dose 2 of the vaccine was offered to all people within the age group (predates the analysis time period for age groups of 60 years and over). B=date on which booster dose was offered to all people within the age group (after the analysis time period for age groups under 60 years).

### Association between of vaccination status and admission rate

The RRR associated with receiving one, two or a booster/three doses of vaccine is shown in Figure 3. For those aged 18–29 years, the RRR associated with two doses increased from 77.4% (95% confidence interval, 49.3% to 91.9%), early in the vaccine roll-out when only those 18–29-year-olds who were clinically extremely vulnerable would have received two doses, to 94.8% (87.8% to 98.2%), which was comparable to that for 30–49-year-olds, by October 2021. Overall, the RRR associated with two doses in 18–29-year-olds was 90.1% (95% CI, 86.8% to 92.7%), for 30–39-year-olds was 92.0% (90.4% to 93.3%), for 40–49-year-olds was 92.5% (91.6% to 93.3%), for 50–59-year-olds was 95.1% (94.7% to 95.5%) and for 60–69-year-olds was 95.9% (95.5% to 96.2%). For those aged 70 years or over, the RRR associated with two doses declined from 97.3% (91.0% to 99.2%) to 86.7% (85.3% to 90.1%) across the study period but increased again to 98.3% (97.6% to 98.8%) with a booster/third dose. The RRR in admission rate for 40–49-year-olds who received a booster/third dose was 92.8% (87.3% to 96.3%), for 50–59-year-olds was 97.7% (96.4% to 98.6%) and for 60–69-year-olds was 97.8% (97.0% to 98.4%). There were not sufficient cases to report on booster/third doses among younger age groups.

**Figure 3.**
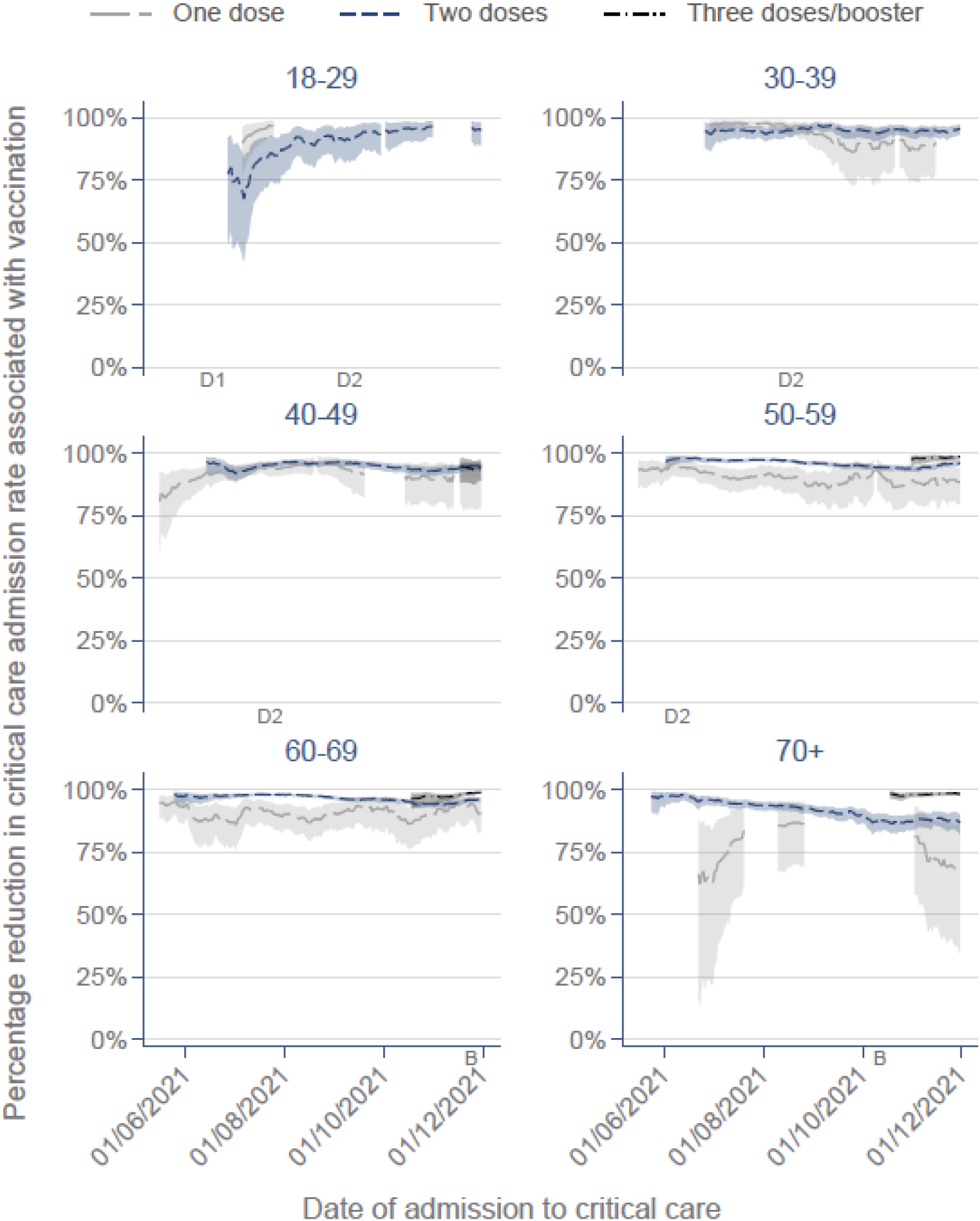
Percentage reduction in rate of admission to critical care associated with vaccination by age group over time. Vaccination status assessed at 14 days prior to the positive SARS-CoV-2 test. 30-day moving average. D1=date on which dose 1 of vaccine was offered to all people within the age group (predates the analysis time period for age groups of 30 years and over). D2=date on which dose 2 of the vaccine was offered to all people within the age group (predates the analysis time period for age groups of 60 years and over). B=date on which booster dose was offered to all people within the age group (after the analysis time period for age groups under 60 years).

### Sensitivity analyses

Assessing vaccination status on the date of the positive SARS-CoV-2 test rather than 14 days prior made negligible difference to the results (Supplementary appendix, Figures S3 and S4).

## Discussion

During the period May to December 2021 in England (when the Delta variant was dominant), the majority of patients admitted for critical care with COVID-19 (59%) were unvaccinated. The characteristics of unvaccinated (compared with vaccinated) patients reflected both vaccination policy (younger and less likely to be immunocompromised) and groups with lower reported vaccine uptake (non-White ethnic groups, those from more deprived areas and, among women, more likely to be pregnant).(22, 23) With the rollout of the vaccination programme, from May to December, the proportion unvaccinated in the general population decreased by three quarters (from 36% to 10%), while the proportion of unvaccinated admitted for critical care decreased by less than one fifth (from 75% to 61%). The impact of vaccination (measured as percentage reduction in rate of admission to critical care) exceeded 92% across all age groups once two doses of vaccine had been rolled out to all adults on 10 September 2021. Additionally, the impact of boosters was also observed, particularly in the oldest age group where some evidence of waning for previous doses was seen.

It is important to note that the observed association between vaccination and lower critical care admission rate may not be entirely causal. For example, higher vaccination uptake is likely to be associated with greater adherence to non-pharmaceutical interventions, such as mask wearing, social distancing, and regular asymptomatic testing.

Our finding of a greater than 92% reduction in critical care admission associated with receiving two doses of vaccination is consistent with two previous observational real-world evaluations of vaccine effectiveness in the United States and in the Netherlands.(11, 13) The study in the United States, conducted when the Alpha variant was the dominant strain, reported vaccine effectiveness of 90% (95% confidence interval 86 to 93%) against critical care admission.(13) The study in the Netherlands reported vaccine effectiveness of 93% (87 to 96%) when Alpha was the dominant strain and 97% (97 to 98%) when Delta was the dominant strain.(11) The results are also similar to estimates of vaccine effectiveness against hospitalisation reported by the UK Health Security Agency of 92% (78 to 97%) for Alpha variant and 94% (85 to 98%) for Delta variant. Our findings contrast somewhat with an Italian retrospective cohort study(24), in which some waning against hospitalisation or death was observed for all age groups. It is possible that hospitalisation and death outcomes include greater proportions of patients who experience the outcome “with” COVID-19 and not “because of” COVID-19, and that the observed waning therefore partly reflected waning against infection and not against severe disease.

The strengths of the study include evaluation of data from the entire of England using data linkage between robust, high-quality datasets. The amount of unlinked data was very low, with only around 1% of vaccination records in NIMS and 1.9% of patients in the ICNARC Case Mix Programme having no NHS number recorded. Population estimates were based on the latest available data from the 2021 census. This is important as even small inaccuracies may lead to substantial imprecision in the size of the denominator for the unvaccinated when vaccine uptake is very high.(25) Vaccination status was assessed at 14 days prior to the positive SARS-CoV-2 test to allow time for vaccination to take effect. The numbers of critical care admissions among recently vaccinated people were too small to be able to evaluate this group as a separate category, and sensitivity analyses demonstrated very similar results if this 14-day lag was not applied.

In conclusion, the vaccination programme in England appears to have substantially reduced admissions for critical care related to COVID-19. While not a causal analysis, our findings are consistent with a substantial and sustained positive impact of vaccination on critical care. For those aged under 70, our findings suggest little waning in protection against critical care admission in the first 6 to 9 months after vaccination.

## Data Availability

The data that support the findings of this study are not publicly available because they are based on de-identified national clinical records. Due to national and organisational data privacy regulations, individual-level data such as those used for this study cannot be shared openly.

## Contributors

DAH, JCD and KMR led the study conceptualisation. All authors contributed to the development of the research question and study design. JH-C led the funding application, obtained data approvals, and undertook the data specification and data linkage. DAH performed the statistical analyses. All authors contributed to the interpretation of the results. DAH, PW, PRM, MS-H and KR wrote the first draft of the paper. All authors contributed to the critical revision of the manuscript for important intellectual content and approved the final version of the manuscript. DAH is guarantor and corresponding author for this work, and attests that all listed authors meet authorship criteria and that no others meeting the criteria have been omitted.

## Acknowledgements

The SGSS data and NIMS data are collated, maintained and quality assured by NHS Digital. NHS Digital bear no responsibility for the analysis or interpretation of the data. We acknowledge the contribution of EMIS Health and the Universities of Nottingham and Oxford for expertise in establishing, developing and supporting the QResearch Trusted Research Environment. This research is part of the Data and Connectivity National Core Study, led by Health Data Research UK in partnership with the Office For National Statistics and funded by UK Research and Innovation (grant ref. MC_PC_20029). MSH is supported by the National Institute for Health Research Clinician Scientist Award (NIHR-CS-2016-16-011). The views expressed in this publication are those of the author(s) and not necessarily those of the NHS, the UK National Institute for Health Research, or the Department of Health.

## References

1. Bravata DM, Perkins AJ, Myers LJ, Arling G, Zhang Y, Zillich AJ, et al. Association of Intensive Care Unit Patient Load and Demand With Mortality Rates in US Department of Veterans Affairs Hospitals During the COVID-19 Pandemic. JAMA Netw Open. 2021;4(1):e2034266.

2. Wilcox ME, Rowan KM, Harrison DA, Doidge JC. Does Unprecedented ICU Capacity Strain, As Experienced During the COVID-19 Pandemic, Impact Patient Outcome? Crit Care Med. 2022;50(6):e548–e56.

3. Zampieri FG, Bastos LSL, Soares M, Salluh JI, Bozza FA. The association of the COVID-19 pandemic and short-term outcomes of non-COVID-19 critically ill patients: an observational cohort study in Brazilian ICUs. Intensive Care Med. 2021;47(12):1440–9.

4. Baraniuk C. Covid-19: How the UK vaccine rollout delivered success, so far. BMJ. 2021;372:421.

5. Intensive Care National Audit and Research Centre (ICNARC). ICNARC report on COVID-19 in critical care, 11 December 2020. Intensive Care National Audit and Research Centre; 2020 11 December 2020.

6. Our World in Data. Number of people vaccinated against COVID-19, Dec 15, 2021 [Available from: https://ourworldindata.org/explorers/coronavirus-data-explorer?zoomToSelection=true&time=2021-12-15&facet=none&pickerSort=asc&pickerMetric=location&Metric=People+vaccinated+%28by+dose%29&Interval=7-day+rolling+average&Relative+to+Population=false&Color+by+test+positivity=false&country=OWID_WRL~GBR.

7. Ferrando-Vivas P, Doidge J, Thomas K, Gould DW, Mouncey P, Shankar-Hari M, et al. Prognostic Factors for 30-Day Mortality in Critically Ill Patients With Coronavirus Disease 2019: An Observational Cohort Study. Crit Care Med. 2021;49(1):102–11.

8. Baden LR, El Sahly HM, Essink B, Kotloff K, Frey S, Novak R, et al. Efficacy and Safety of the mRNA-1273 SARS-CoV-2 Vaccine. N Engl J Med. 2021;384(5):403–16.

9. Falsey AR, Sobieszczyk ME, Hirsch I, Sproule S, Robb ML, Corey L, et al. Phase 3 Safety and Efficacy of AZD1222 (ChAdOx1 nCoV-19) Covid-19 Vaccine. N Engl J Med. 2021;385(25):2348–60.

10. Polack FP, Thomas SJ, Kitchin N, Absalon J, Gurtman A, Lockhart S, et al. Safety and Efficacy of the BNT162b2 mRNA Covid-19 Vaccine. N Engl J Med. 2020;383(27):2603–15.

11. de Gier B, Kooijman M, Kemmeren J, de Keizer N, Dongelmans D, van Iersel SCJL, et al. COVID-19 vaccine effectiveness against hospitalizations and ICU admissions in the Netherlands, April-August 2021. medRxiv. 2021:2021.09.15.21263613.

12. Hilty MP, Keiser S, Wendel Garcia PD, Moser A, Schuepbach RA. mRNA-based SARS-CoV-2 vaccination is associated with reduced ICU admission rate and disease severity in critically ill COVID-19 patients treated in Switzerland. Intensive Care Med. 2022;48(3):362–5.

13. Thompson MG, Stenehjem E, Grannis S, Ball SW, Naleway AL, Ong TC, et al. Effectiveness of Covid-19 Vaccines in Ambulatory and Inpatient Care Settings. N Engl J Med. 2021;385(15):1355–71.

14. Lintern S. Doctors and nurses vent anger as unvaccinated Covid cases delay vital operations. The Times. 2021 4 December 2021.

15. Yan H, Elamroussi A. Unvaccinated Covid-19 patients are filling up hospitals, putting the care of others at risk, doctors say. CNN. 2021 1 August 2021.

16. Health Data Research Innovation Gateway. Covid-19 Second Generation Surveillance System [Available from: https://web.www.healthdatagateway.org/dataset/b15760f3-e188-446f-aee5-c38bd3859dd4.

17. Health Data Research Innovation Gateway. National Immunisation Management System (NIMS) [Available from: https://web.www.healthdatagateway.org/dataset/65b04cbe-1b5c-44d7-b640-ce63fb106401.

18. GOV.UK Coronavirus (COVID-19) in the UK [Available from: https://coronavirus.data.gov.uk/details/download.

19. Office for National Statistics. First results from Census 2021 in England and Wales [Available from: https://www.ons.gov.uk/releases/initialfindingsfromthe2021censusinenglandandwales.

20. Lopez Bernal J, Andrews N, Gower C, Robertson C, Stowe J, Tessier E, et al. Effectiveness of the Pfizer-BioNTech and Oxford-AstraZeneca vaccines on covid-19 related symptoms, hospital admissions, and mortality in older adults in England: test negative case-control study. BMJ. 2021;373:1088.

21. Rothman KJ. Modern Epidemiology. Boston: Little, Brown; 1986.

22. Curtis HJ, Inglesby P, Morton CE, MacKenna B, Green A, Hulme W, et al. Trends and clinical characteristics of COVID-19 vaccine recipients: a federated analysis of 57.9 million patients’ primary care records in situ using OpenSAFELY. Br J Gen Pract. 2022;72(714):e51–e62.

23. Iacobucci G. Covid-19 and pregnancy: vaccine hesitancy and how to overcome it. BMJ. 2021;375:2862.

24. Fabiani M, Puopolo M, Morciano C, Spuri M, Spila Alegiani S, Filia A, et al. Effectiveness of mRNA vaccines and waning of protection against SARS-CoV-2 infection and severe covid-19 during predominant circulation of the delta variant in Italy: retrospective cohort study. BMJ. 2022;376:e069052.

25. Groyer A. COVID-19 Actuaries Response Group. 2021. [1 February 2022]. Available from: https://covidactuaries.org/2021/10/27/vaccine-effectiveness-and-population-estimates/.

