## Supplementary appendix for "Impact of vaccination on COVID-19-associated admissions to critical care in England: a population cohort study of linked data"

David A Harrison, Peter J Watkinson, James C Doidge, Manu Shankar-Hari, Paul R Mouncey, Martina Patone, Carol A C Coupland, Julia Hippisley-Cox, Kathryn M Rowan

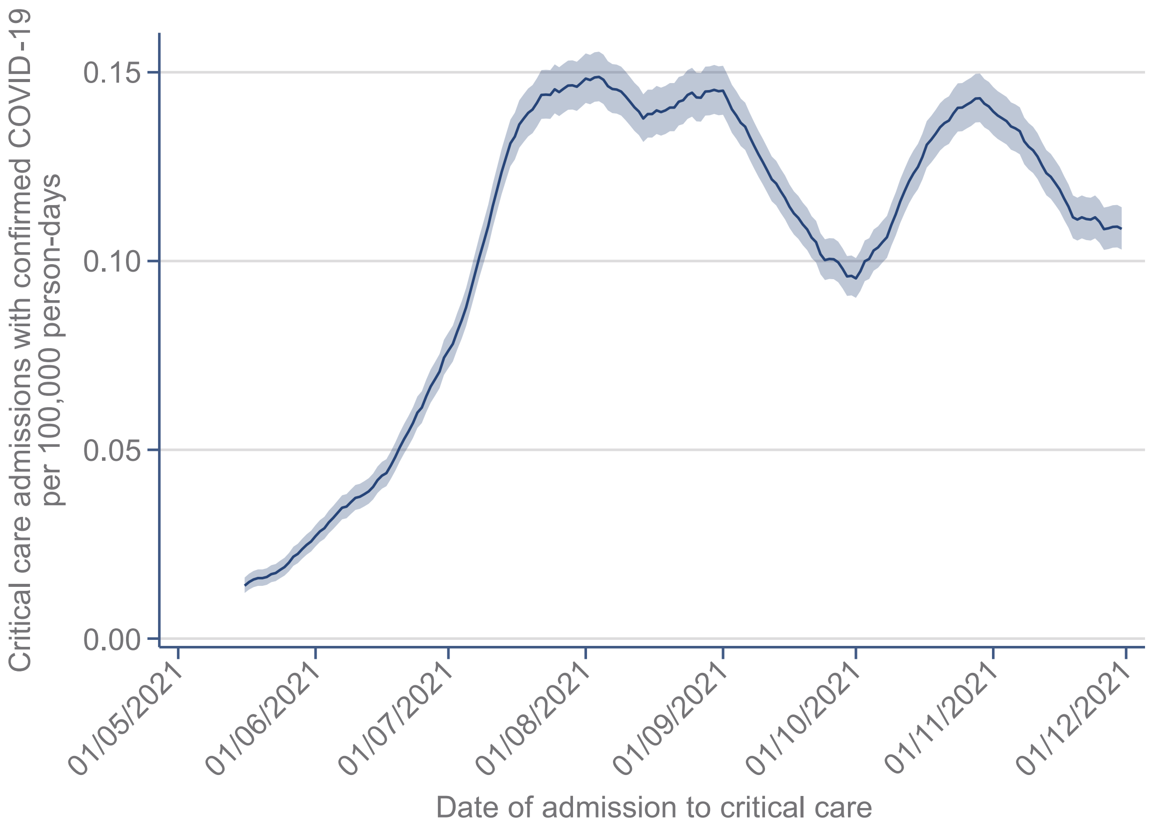

**Figure S1. Rate of admission to critical care over time**
30-day moving average.

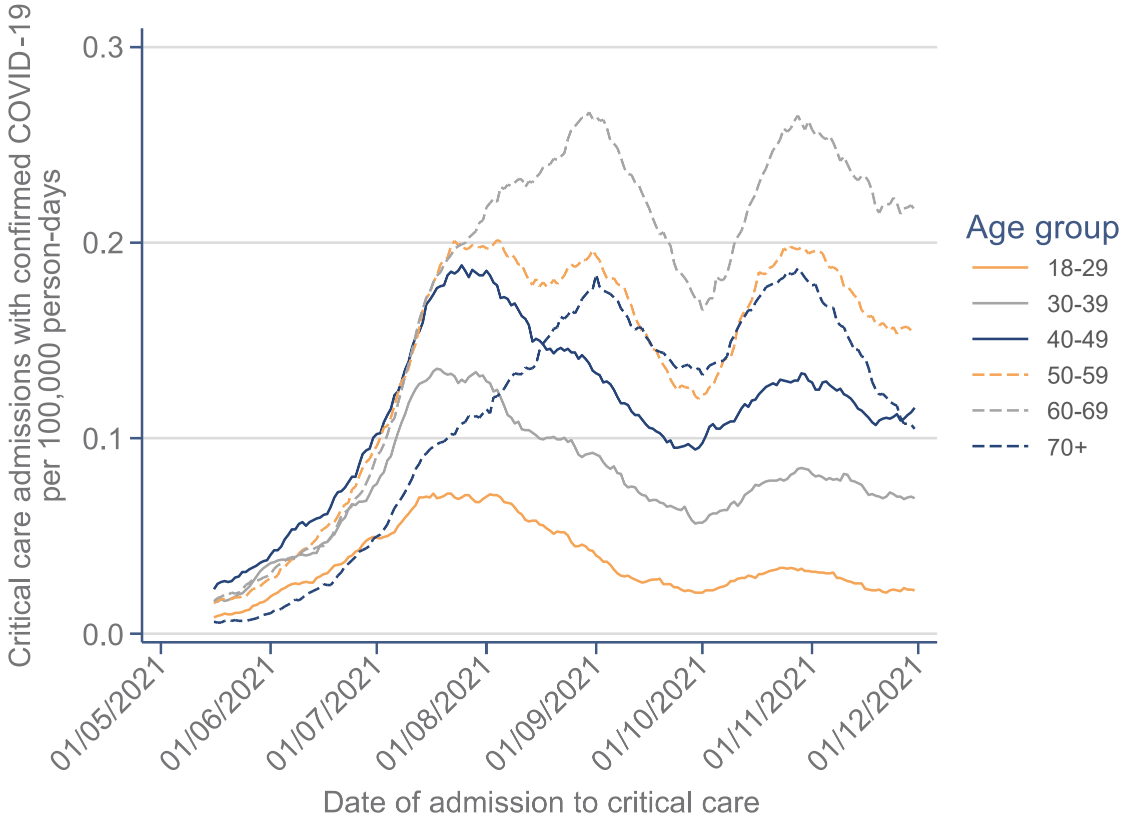

**Figure S2. Rate of admission to critical care by age group over time**
30-day moving average.

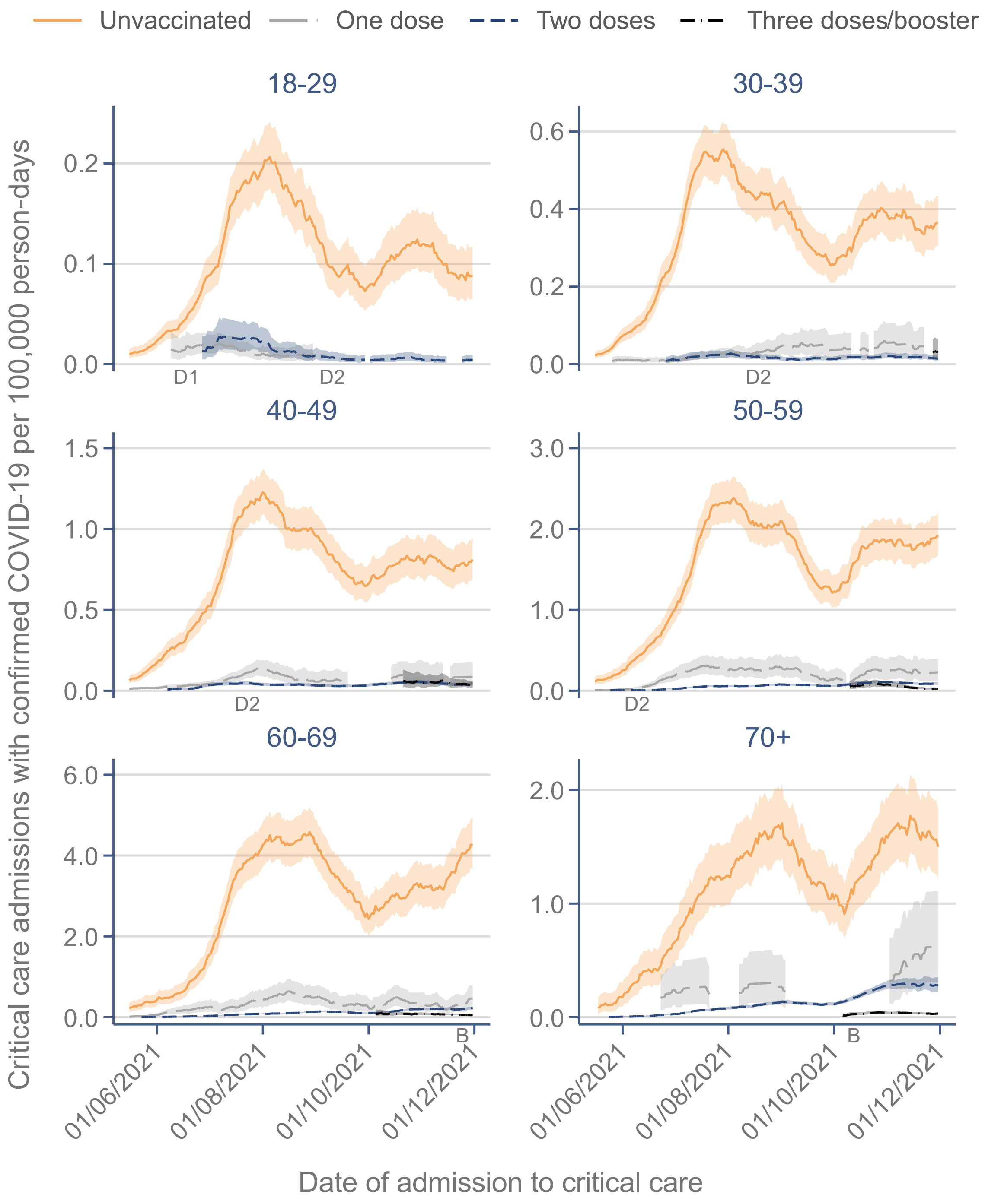

**Figure S3. Rate of admission to critical care by age group and vaccination status over time – sensitivity analysis**

Vaccination status assessed on the date of the positive SARS-CoV-2 test. 30-day moving average. D1=date on which dose 1 of vaccine was offered to all people within the age group (predates the analysis time period for age groups of 30 years and over). D2=date on which dose 2 of the vaccine was offered to all people within the age group (predates the analysis time period for age groups of 60 years and over). B=date on which booster dose was offered to all people within the age group (after the analysis time period for age groups under 60 years).

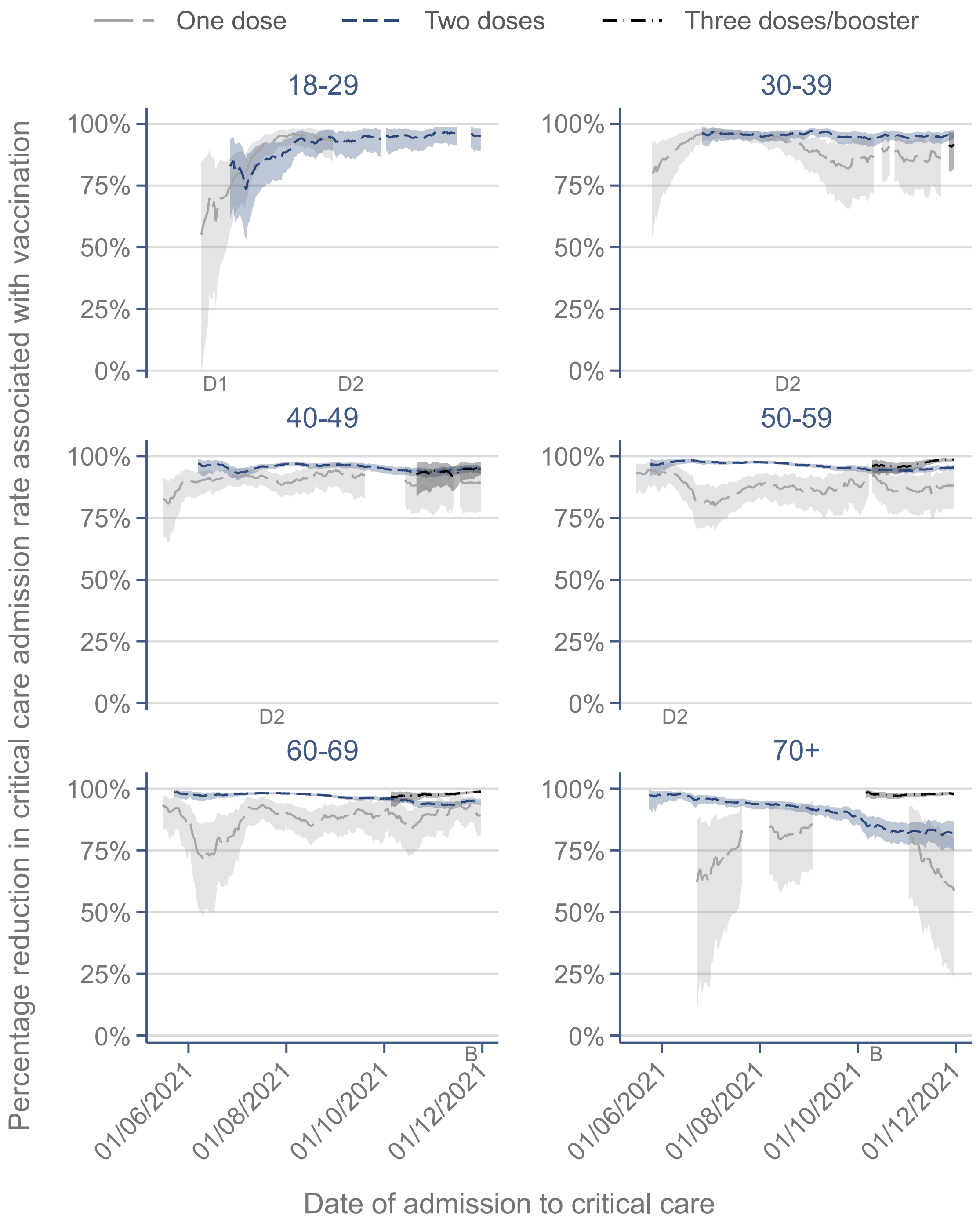

**Figure S4. Percentage reduction in rate of admission to critical care associated with vaccination by age group over time – sensitivity analysis**
Vaccination status assessed on the date of the positive SARS-CoV-2 test. 30-day moving average. D1=date on which dose 1 of vaccine was offered to all people within the age group (predates the analysis time period for age groups of 30 years and over). D2=date on which dose 2 of the vaccine was offered to all people within the age group (predates the analysis time period for age groups of 60 years and over). B=date on which booster dose was offered to all people within the age group (after the analysis time period for age groups under 60 years).
